# Patient-derived PixelPrint phantoms for evaluating clinical imaging performance of a deep learning CT reconstruction algorithm

**DOI:** 10.1101/2023.12.07.23299625

**Authors:** Jessica Y. Im, Sandra S. Halliburton, Kai Mei, Amy E. Perkins, Eddy Wong, Leonid Roshkovan, Olivia F. Sandvold, Leening P. Liu, Grace J. Gang, Peter B. Noël

**Affiliations:** Department of Radiology, University of Pennsylvania, Philadelphia, PA, USA; Department of Bioengineering, University of Pennsylvania, Philadelphia, PA, USA; Philips Healthcare, Orange Village, OH, USA

**Keywords:** Deep learning reconstruction (DLR), CT imaging phantoms, 3D-printed imaging phantoms, Dose reduction

## Abstract

**Objective:** Deep learning reconstruction (DLR) algorithms exhibit object-dependent resolution and noise performance. Thus, traditional geometric CT phantoms cannot fully capture the clinical imaging performance of DLR. This study uses a patient-derived 3D-printed PixelPrint lung phantom to evaluate a commercial DLR algorithm across a wide range of radiation dose levels.

**Approach:** The lung phantom used in this study is based on a patient chest CT scan containing ground glass opacities and was fabricated using PixelPrint 3D-printing technology. The phantom was placed inside two different sized extension rings to mimic a small and medium sized patient and was scanned on a conventional CT scanner at exposures between 0.5 and 20 mGy. Each scan was reconstructed using filtered back projection (FBP), iterative reconstruction, and DLR at five levels of denoising. Image noise, contrast to noise ratio (CNR), root mean squared error (RMSE), structural similarity index (SSIM), and multi-scale SSIM (MS SSIM) were calculated for each image.

**Main Results:** DLR demonstrated superior performance compared to FBP and iterative reconstruction for all measured metrics in both phantom sizes, with better performance for more aggressive denoising levels. DLR was estimated to reduce dose by 25-83% in the small phantom and by 50-83% in the medium phantom without decreasing image quality for any of the metrics measured in this study. These dose reduction estimates are more conservative compared to the estimates obtained when only considering noise and CNR with a non-anatomical physics phantom.

**Significance:** DLR has the capability of producing diagnostic image quality at up to 83% lower radiation dose which can improve the clinical utility and viability of lower dose CT scans. Furthermore, the PixelPrint phantom used in this study offers an improved testing environment with more realistic tissue structures compared to traditional CT phantoms, allowing for structure-based image quality evaluation beyond noise and contrast-based assessments.

## 1. Introduction

Over the last few years, there has been substantial interest in the development and clinical use of deep learning reconstruction (DLR) algorithms for improving computed tomography (CT) image quality and reducing radiation dose^1^. For decades, filtered back projection (FBP) was the dominant reconstruction algorithm due to its numerical stability and fast computation time^2^. However, at lower doses, FBP image quality drops and image noise increases dramatically^1^. With continued interest in dose reduction^3^, especially in pediatric populations^4–7^, clinical CT imaging has begun moving away from FBP toward newer solutions such as iterative reconstruction (IR) which preserves image quality at lower doses. Various forms of IR have demonstrated significant potential to minimize noise and thus to reduce dose compared to FBP^8^. However, limitations in IR including unnatural noise texture^8,9^ and extended reconstruction time^1,8^ have resulted in a push for further innovation in reconstruction solutions.

DLR for CT has emerged as a novel solution for improving image quality and reconstruction time while preserving FBP-like noise textures. These algorithms utilize artificial neural networks such as convolutional neural networks (CNNs)^10,11^ or generative adversarial networks (GANs)^12^ which are trained to produce optimized output images from lower dose input data. DLR frameworks can be broadly categorized as either indirect, where a deep learning network is used alongside FBP or IR, or direct, in which the network directly converts sinogram data to image data without FBP or IR^1^. Many different implementations of DLR have been proposed in academic research^13–15^ as well as introduced clinically by CT vendors^9,16,17^.

With the rise of commercially available DLR algorithms, there has been an increase in studies evaluating DLR. Multiple patient and phantom studies have demonstrated that DLR can improve image quality at low doses through enhanced lesion detectability and reduced noise^5–7,18–25^. These studies utilize quantitative metrics such as signal-to-noise ratio (SNR), contrast-to-noise ratio (CNR), noise, detectability index (d’), and noise power spectrum (NPS). In addition, qualitative scores for various aspects of subjective image quality have been obtained via reader studies by experienced radiologists. The literature has shown that various implementations of DLR can reduce dose by about 30-71% compared to hybrid iterative reconstruction (HIR) methods while preserving diagnostic image quality^1^.

While there are many promising results regarding DLR performance, there are several limitations to current studies. First, due to the nonlinear nature of DLR, images reconstructed with DLR demonstrate object-dependent resolution and noise^26^. Traditional CT phantoms used in DLR evaluation studies are often composed of simple geometric shapes which are not designed to represent realistic tissue structures^19,22,23^. As a result, general image quality metrics such as noise and CNR measured on traditional CT phantoms cannot fully capture the clinical imaging performance of DLR. Second, clinical imaging studies using patient data are often limited by sample size and restricted by radiation dose exposure concerns^24,25,27^, which limit the acceptable dose range as well as the number of times a patient can be scanned. Furthermore, patient scans do not have reliable ground truth images for comparison and thus cannot be used to assess the structural accuracy of a reconstructed image. A clinical scenario in which structural accuracy is important is lung CT imaging with ground glass opacity (GGO) findings. Subtle differences in shape (round vs polygonal, with or without radial growths) and texture (presence or absence of solid densities) in a GGO can lead to differences in image interpretation and clinical decision making^28^. Because of this, the accurate reconstruction of such structures and details is critical to ensuring the highest quality of patient care. Image reconstruction for clinical scenarios such as this require evaluation beyond what is available with current phantom and patient studies.

This study proposes to use a patient-derived PixelPrint^29–31^ phantom as a novel solution to address the current limitations in the evaluation of DLR performance. PixelPrint is a technology which produces 3D-printed patient-based phantoms which demonstrate highly detailed tissue structures, realistic textures, and accurate attenuation profiles. PixelPrint software converts 3D CT images into geometric code (g-code) instructions for fused filament fabrication (FFF) 3D printers by taking advantage of the partial volume effect to produce desired Hounsfield Unit (HU) values^31^. Previous studies have demonstrated a high degree of HU and geometric similarity between scans of PixelPrint phantoms and their reference patient scans^32^. Furthermore, reader studies demonstrated that there was no clinically significant difference in image quality assessment between reading a phantom lung image and reading a patient lung image^30^. Compared to standard geometric CT imaging phantoms, PixelPrint phantoms demonstrate realistic tissue morphology and thus can more fully capture the clinical imaging performance of DLR. Compared to patient data, PixelPrint phantoms allow for more flexibility in radiation dose usage and have more accurate ground truth images with which to assess the structural precision of DLR images.

This study utilized a 3D-printed PixelPrint lung phantom to evaluate the clinical imaging performance of a commercial DLR algorithm, Precise Image (PI) (Philips Healthcare, Cleveland, OH, USA)^9^, in comparison to FBP and IR. PI is an example of a direct DLR algorithm and utilizes simulated low dose sinogram data for CNN training^1,9^.The PixelPrint phantom was scanned with a large range of radiation doses to investigate the dose reduction potential of each algorithm. As image quality is affected by patient size (i.e., CT images of large patients tend to have higher noise and reduced image quality compared to smaller patients), two different phantom sizes were included in the performance assessment to examine the generalizability of results to different patient sizes.

## 2. Methods

### 2.1 Patient CT Scan Selection

A single patient chest CT scan containing multiple subsolid GGOs representing metastatic lesions was retrospectively selected as the model for the 3D-printed phantom in this study (Figure 1). GGO lesions are an example of highly detailed lung structures in which accurate reconstruction of textures and shapes is clinically important. The Institutional Review Board (IRB) at the University of Pennsylvania approved this retrospective study, and the image was taken from the Hospital of the University of Pennsylvania PACS system and anonymized. The scan and reconstruction parameters of the patient CT scan are listed in Table 1.

**Figure 1.**
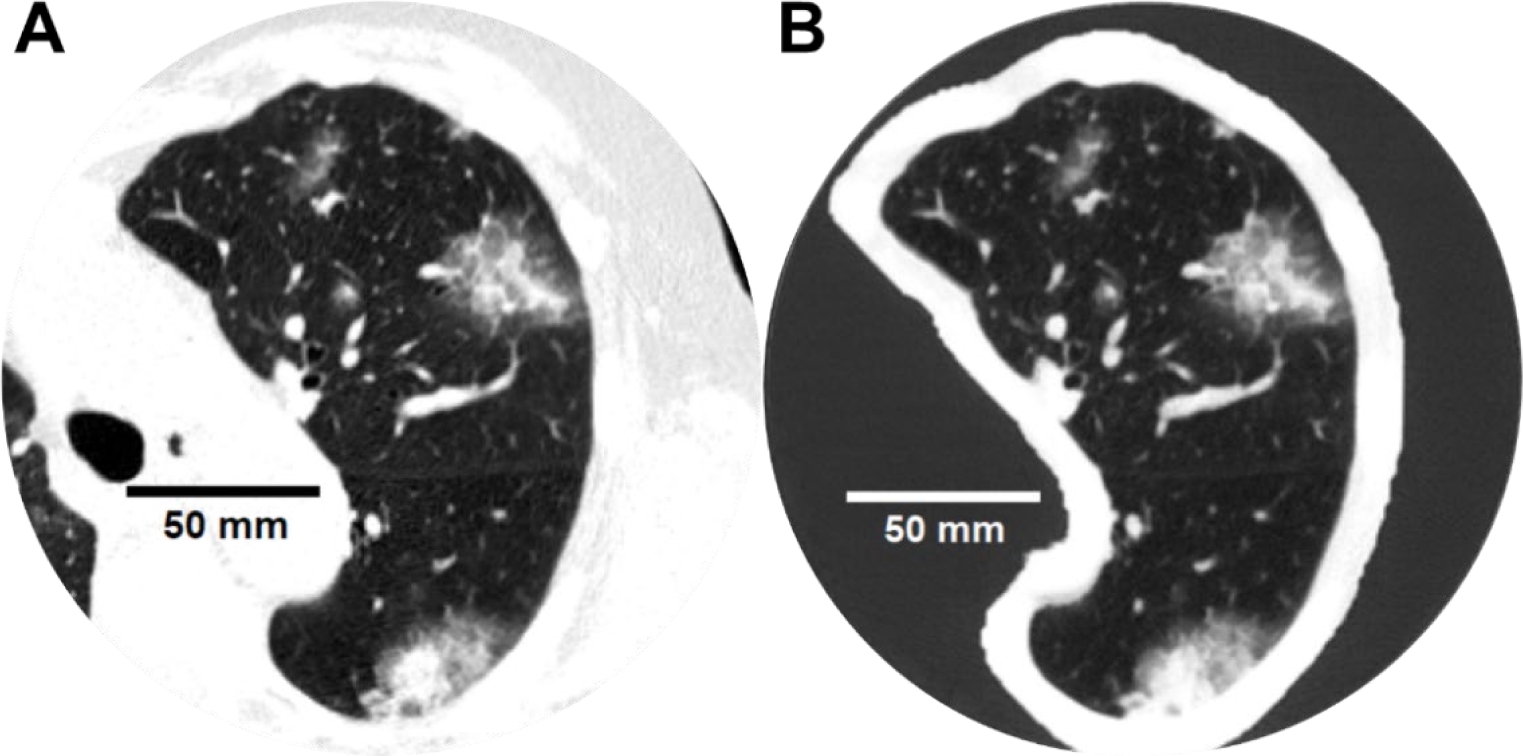
(A) Image of the left lung from the patient chest CT scan which was used to generate the phantom. (B) CT scan of the printed phantom, scanned at 20 mGy and 120 kVp and reconstructed using PI-Sharp. WL: −500, WW: 1000

**Table 1.** Scan and reconstruction parameters of the patient CT image.

| Scanner Model | Philips Spectral CT 7500 |
| --- | --- |
| Scan mode | Helical |
| Tube voltage | 120 kVp |
| Tube current | 173 mA |
| Rotation time | 0.4 s |
| Helical pitch | 1.15 |
| Exposure | 60 mAs |
| CTDI<sub>vol</sub> | 4.7 mGy |
| Collimation | 128 x 0.625 mm |
| Slice thickness | 1 mm |
| Slice increment | 1 mm |
| Reconstruction filter | YA |
| Reconstructed field of view | 368 x 368 mm<sup>2</sup> |
| Matrix size | 512 x 512 pixel<sup>2</sup> |
| Pixel spacing | 0.7188 mm |

### 2.2 Phantom Fabrication

The phantom was fabricated using PixelPrint technology^30,31^ to produce a realistic patient-specific lung CT phantom. The phantom was designed as a 20 cm diameter cylinder containing the segmented left lung positioned in the center of the cylinder. A 4 cm scan length containing a large (4.5 x 3.2 cm) GGO was selected. The left lung was segmented along with a 1 cm border of surrounding tissue using an open-source automated U-net lung segmentation model^33^. The regions inside of the segmented lung were printed using PixelPrint technology to modulate density and accurately reproduce the HU profiles of the patient image, while regions of the cylinder surrounding the segmented anatomy were printed with a constant infill ratio of 15% (corresponding to ∼-800 HU). The entire phantom was 3D-printed as one piece using polylactic acid (PLA) filament on an FFF printer (Lulzbot TAZ Sidekick with M175 v2 tool head, Fargo Additive Manufacturing Equipment 3D, LLC Fargo, ND, USA).

### 2.3 Image Acquisition and Reconstruction

The phantom was scanned with a default high resolution chest imaging protocol on a conventional CT scanner (Incisive CT, Philips Healthcare, Cleveland, OH, USA). Multiple scans were acquired with varying radiation dose levels ranging from 0.5 to 20 mGy. Scans were repeated three times at each dose level and each scan was reconstructed using FBP, an iterative reconstruction algorithm (iDose^4^) at a single noise level (Level 3), and DLR (Precise Image (PI)) at five levels with increasingly aggressive noise reduction (Sharper, Sharp, Standard, Smooth, Smoother) (Table 2). Additional scan and reconstructions parameters common to all scans are listed in Table 3.

**Table 2.** Varying radiation dose levels used for phantom scanning and the different methods used for reconstruction.

| Exposure [mAs] | CTDI<sub>vol</sub> [mGy] | Reconstruction Algorithms |
| --- | --- | --- |
| 250* | 20* | FBP, YC Filter*<br>iDose<sup>4</sup> Level 3, YC filter<br>Precise Image (PI), Lung Definition, Sharper/Sharp/Standard/Smooth/Smooother |
| 235 | 19 |  |
| 185 | 15 |  |
| 148<sup>†</sup> | 12<sup>†</sup> |  |
| 111<sup>‡</sup> | 9<sup>‡</sup> |  |
| 74 | 6 |  |
| 49 | 4 |  |
| 25<sup>§</sup> | 2<sup>§</sup> |  |
| 12 | 1 |  |
| 6 | 0.5 |  |

\* Dose and reconstruction parameters used for ground truth image.
<sup>†</sup> Diagnostic reference level for a 29-33cm water-equivalent diameter patient.<sup>34</sup>
<sup>‡</sup> Achievable dose level for a 29-33cm water-equivalent diameter patient.<sup>34</sup>
<sup>§</sup> Lung cancer screening level.<sup>35</sup>

**Table 3.** CT scan and reconstruction parameters for the phantom scans.

| Scanner Model | Philips Incisive CT |
| --- | --- |
| Scan mode | Helical |
| Tube voltage | 120 kVp |
| Rotation time | 0.5 s |
| Helical pitch | 1 |
| Collimation | 64 x 0.625 mm |
| Slice thickness | 1 mm |
| Slice increment | 0.5 mm |
| Reconstructed field of view | 350 x 350 mm<sup>2</sup> |
| Matrix size | 512 x 512 pixel<sup>2</sup> |
| Pixel spacing | 0.6836 mm |

### 2.4 Extension Rings

To mimic different patient sizes, the phantom was placed inside two different sized extension rings during scanning (Figure 2). A custom 25 x 25 cm water-equivalent extension ring with a 20 cm cylindrical bore was 3D printed using PLA filament. The phantom was placed in this custom extension ring to represent a small sized patient (small phantom), resulting in a total water equivalent diameter of about 19 cm. For a medium size, the phantom was placed in the 20 cm bore of a 30 x 40 cm multi-energy CT phantom (MECT) (Sun Nuclear, WI, USA) extension ring (medium phantom). The total water equivalent diameter of the phantom plus MECT extension ring was about 30 cm. The scan and reconstruction parameters outlined in Tables 2 and 3 were repeated for each phantom size.

**Figure 2.**
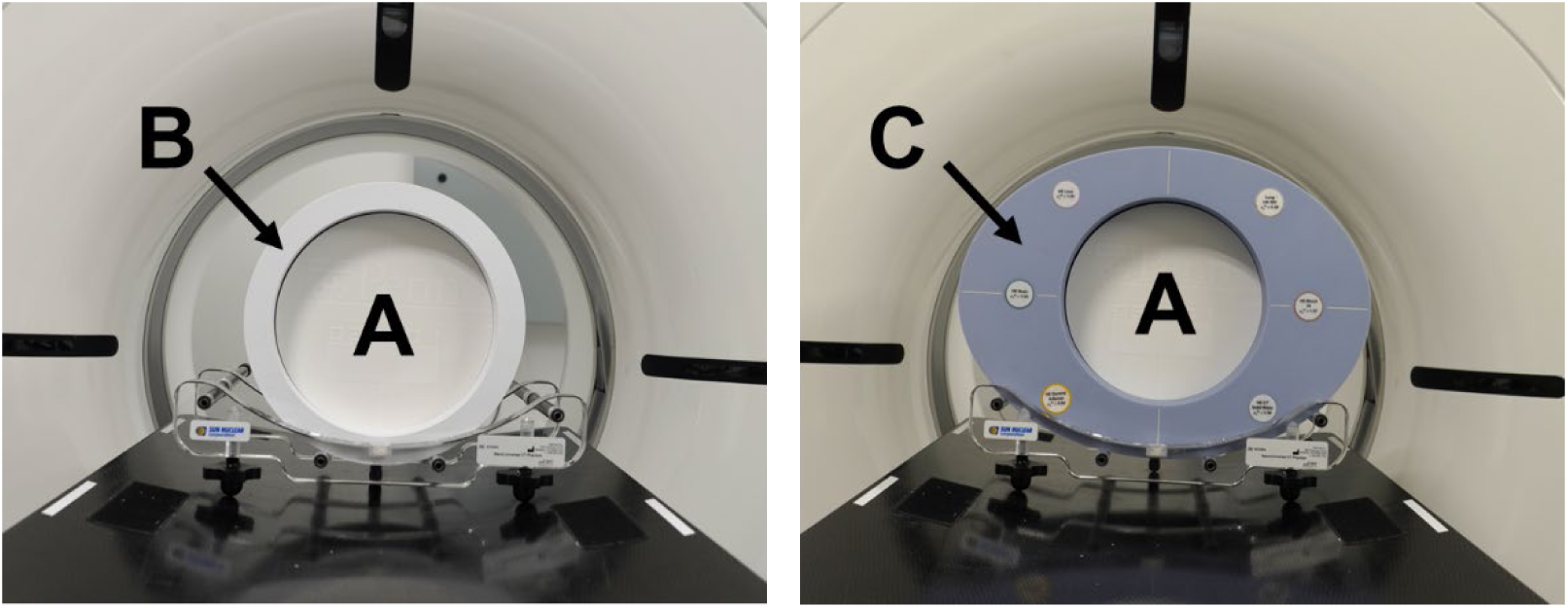
The PixelPrint lung phantom (A) placed inside of a 25 x 25 cm 3D printed extension ring (B) to represent a small sized pateient and placed inside of a 30 x 40 cm MECT extension ring (C) to represent a medium sized patient.

### 2.5 Image Analysis

Image noise and CNR were calculated for each reconstruction and dose combination. The image noise was calculated for a 2 x 2 cm region of interest (ROI) across 10 consecutive slices in a homogeneous region of the phantom background lung parenchyma. The CNR was calculated between the GGO lesion and the background lung parenchyma where the GGO ROI was a 2 x 2 cm ROI over 14 consecutive slices inside of the GGO lesion. The equations used for noise and CNR calculations were:

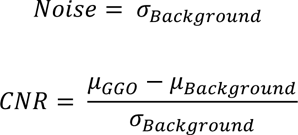

Where 𝜎*_Background_* is the standard deviation of HU values in the background lung ROI, 𝜇*_GGo_* is the mean HU in the GGO ROI, and 𝜇*_Background_* is the mean HU in the background lung ROI.

In addition to these general image quality metrics, structural accuracy of the reconstructed images was evaluated using the image similarity metrics: root mean squared error (RMSE), structural similarity index measure (SSIM)^36^, and multi-scale SSIM (MS SSIM)^37^. These metrics were measured in a 13.5 x 13.5 cm ROI across 50 consecutive slices within the 3D printed phantom, using the highest dose (20 mGy) FBP image as the ground truth image. Since the 20 mGy scans were used as the ground truth, they were excluded from the sample for all image metric calculations. The RMSE and SSIM were calculated for each image using the open source Python package skimage.metrics^38^, and the MS SSIM was calculated using the open source python library pytorch-msssim^39^. All ROIs used for these calculations are shown in Figure 3, and the same ROIs were used for each of the reconstructed images.

**Figure 3.**
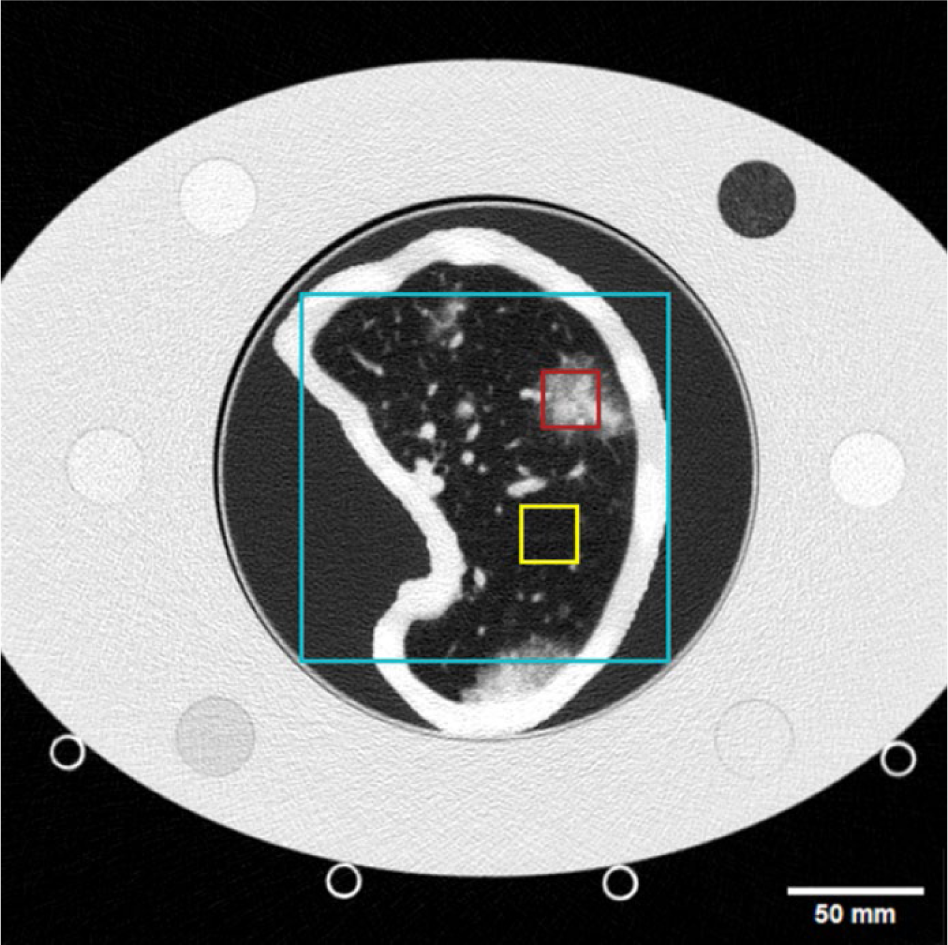
CT scan of the phantom placed inside of the medium sized extension ring with marked ROIs used for image quality metric measurements. The yellow box encompasses the background lung ROI used for image noise and CNR calculations, the red box shows the GGO ROI used to calculate CNR, and the cyan box represents the ROI used for RMSE, SSIM, and MS SSIM measurements. WL: −450, WW: 1100.

### 2.6 Statistical analysis

The performance of each dose and reconstruction combination was evaluated in comparison to the performance of the FBP images of scans taken at 12 mGy, which is the diagnostic reference level for 29-33 cm water equivalent diameter patients^34^. A two-sample, one-tailed t-test was performed for each image metric using the open-source python package Scipy statistical functions^40^. Effects were considered statistically significant where 𝑝 < 0.05, which after applying the Bonferroni *post hoc* correction results in 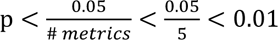. The potential dose reduction of each reconstruction algorithm was then determined by finding the lowest dose measured at which there was no statistically significant decrease in image quality from the reference for any measured metric.

## 3. Results

### 3.1 Comparison of reconstruction algorithms

PI demonstrates superior performance compared to both FBP and iterative reconstruction for all measured metrics in both phantom sizes. The image quality of FBP images is noticeably degraded by noise at lower doses while low dose scans reconstructed with iDose^4^ and PI have image quality which more closely resemble the highest dose FBP image. This effect is demonstrated visually in Figure 4 and confirmed quantitatively by the measured metrics. The results of each metric are represented in Figures 5 and 6, and Tables A1-A10 show the *t*-test statistics.

**Figure 4.**
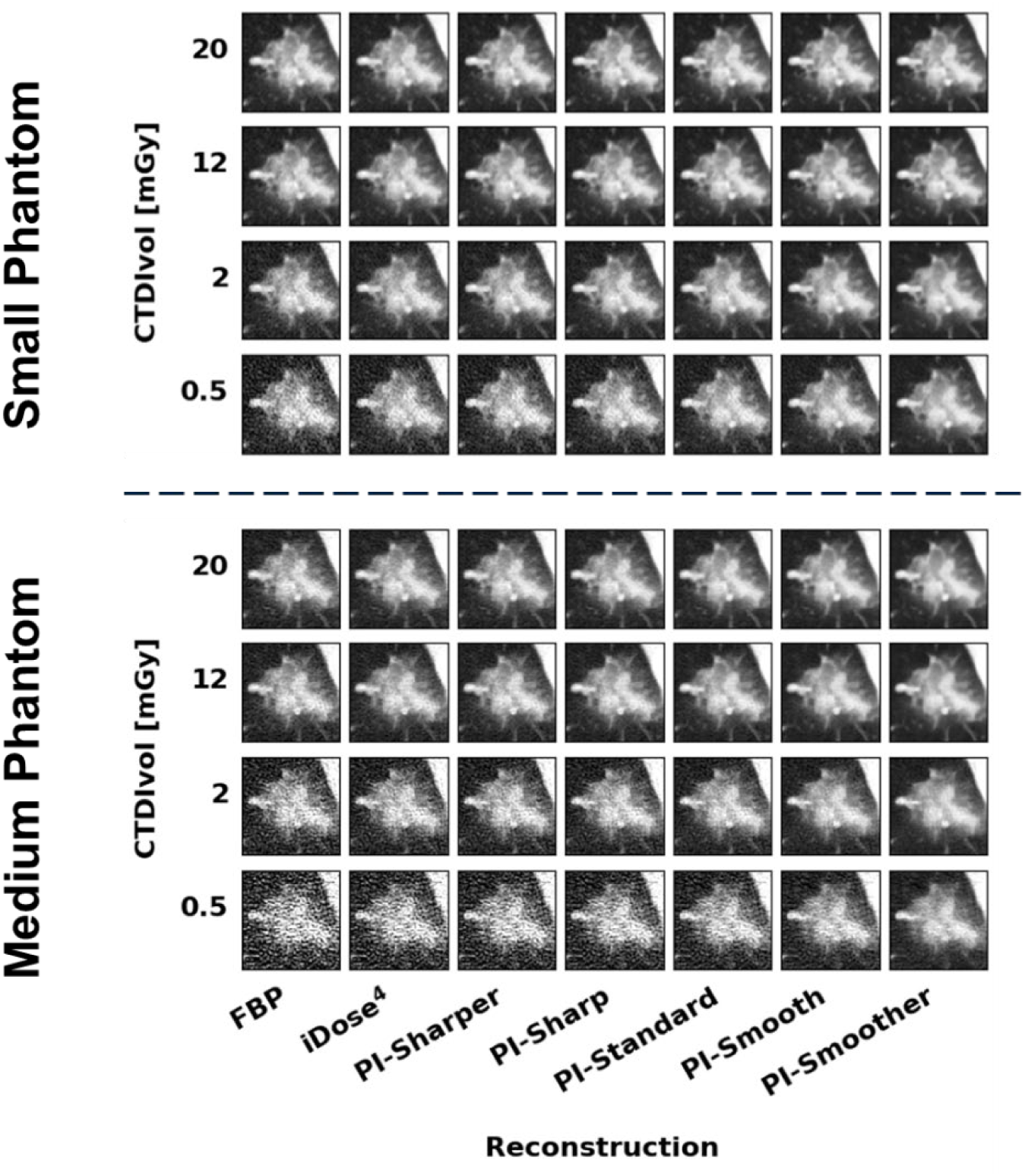
Images of the GGO lesion taken from the small phantom (top) and medium phantom (bottom) at several dose and reconstruction combinations. Figures A1 and A2 in the appendix show the GGO lesion images from all the dose levels collected. WL: −500, WW: 1000.

**Figure 5.**
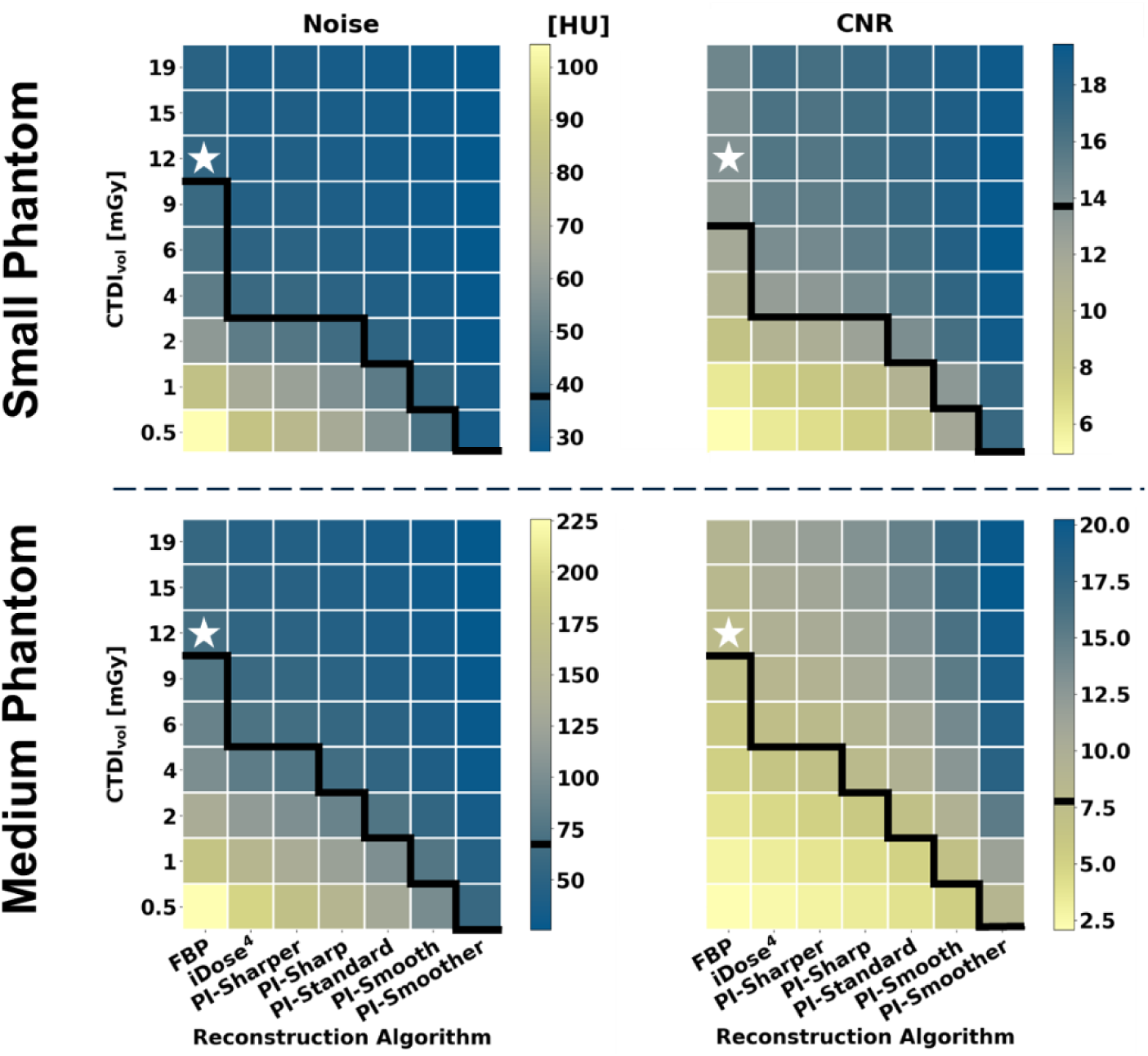
Heatmaps displaying results of the image metric calculations for Noise (left) and CNR (right) from both the small phantom (top) and medium phantom (bottom). A white star is used to designate the dose and reconstruction combination used as the reference for statistical comparison for each metric. The value for this reference group is indicated on the color bars by a black line. The black lines on the heatmaps separate values that are statistically better than or equivalent to the reference (above the line) from those that are statistically worse than the reference (below the line).

**Figure 6.**
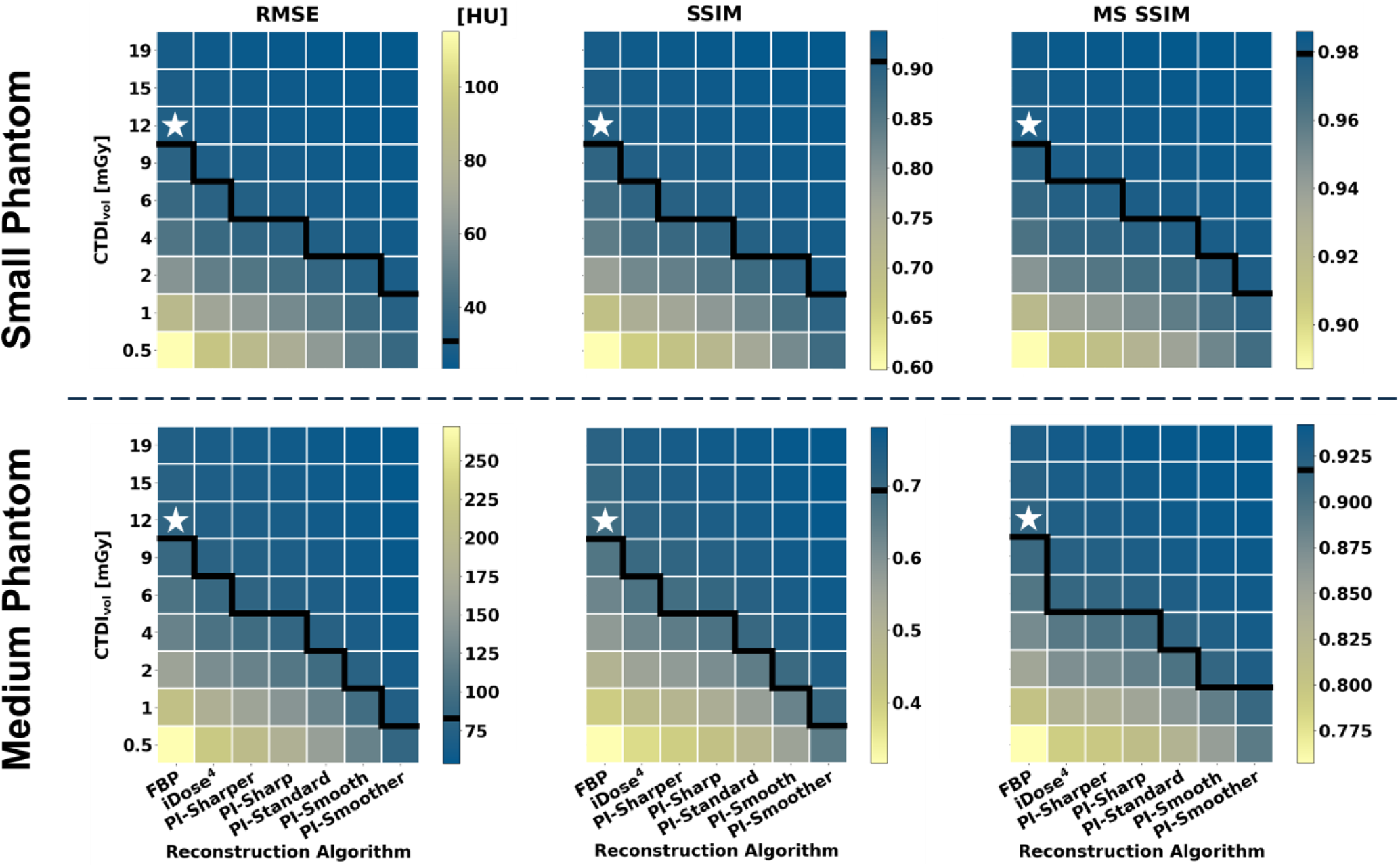
Heatmaps displaying results of the image metric calculations for RMSE (left), SSIM (middle), and MS SSIM (right) from both the small phantom (top) and medium phantom (bottom). The white stars and black lines in this figure have the same function as in Figure 5.

All metrics show that iDose^4^ is capable of dose reduction compared to FBP and that PI shows further dose reduction compared to iDose^4^. Furthermore, more aggressive noise reduction, i.e. smoother levels of PI, showed improved performance over less aggressive noise reduction, i.e. sharper levels of PI. When only considering noise and CNR, the different levels of PI achieved dose reduction capabilities between 67-96% for the small phantom and between 50-96% for the medium phantom, respectively (Figure 5). However, the results of the image similarity metrics RMSE, SSIM, and MSSIM show more conservative dose reduction estimates compared to the estimates obtained from noise and CNR alone. When considering the image similarity metric results, PI demonstrated lower dose reduction capabilities of 25-83% in the small phantom and 50-83% in the medium phantom (Figure 6). Thus, these image similarity metrics provide additional information about the structural accuracy of the reconstructed images that is not captured by general image quality metrics like noise and CNR.

### 3.2 Phantom Size Effects

The image quality of the small phantom reconstructions showed an average of approximately 40% improvement across all metrics compared to the matched doses and reconstructions of the medium phantom. Analysis of noise and CNR suggest that there is a slight increase in dose reduction capabilities of PI in the small phantom (67-96%) compared to the medium phantom (50-96%). However, the image similarity metrics show the opposite trend, with slightly lower dose reduction capabilities in the small phantom (25-83%) compared with the medium phantom (50-83%). Overall the results from the two phantom sizes showed similar trends in dose reduction.

### 3.3 Summarized Potential Dose Reduction Capabilities

The overall dose reduction of each reconstruction algorithm compared to FBP was determined using the minimum dose at which all metrics matched or exceeded the reference performance. These minimum doses are summarized in Figure 7 with corresponding dose reduction percentages indicated on the right axis. For both the small phantom and medium phantom, PI demonstrated dose reduction capabilities up to 83% for the highest level of denoising (PI-Smoother).

**Figure 7.**
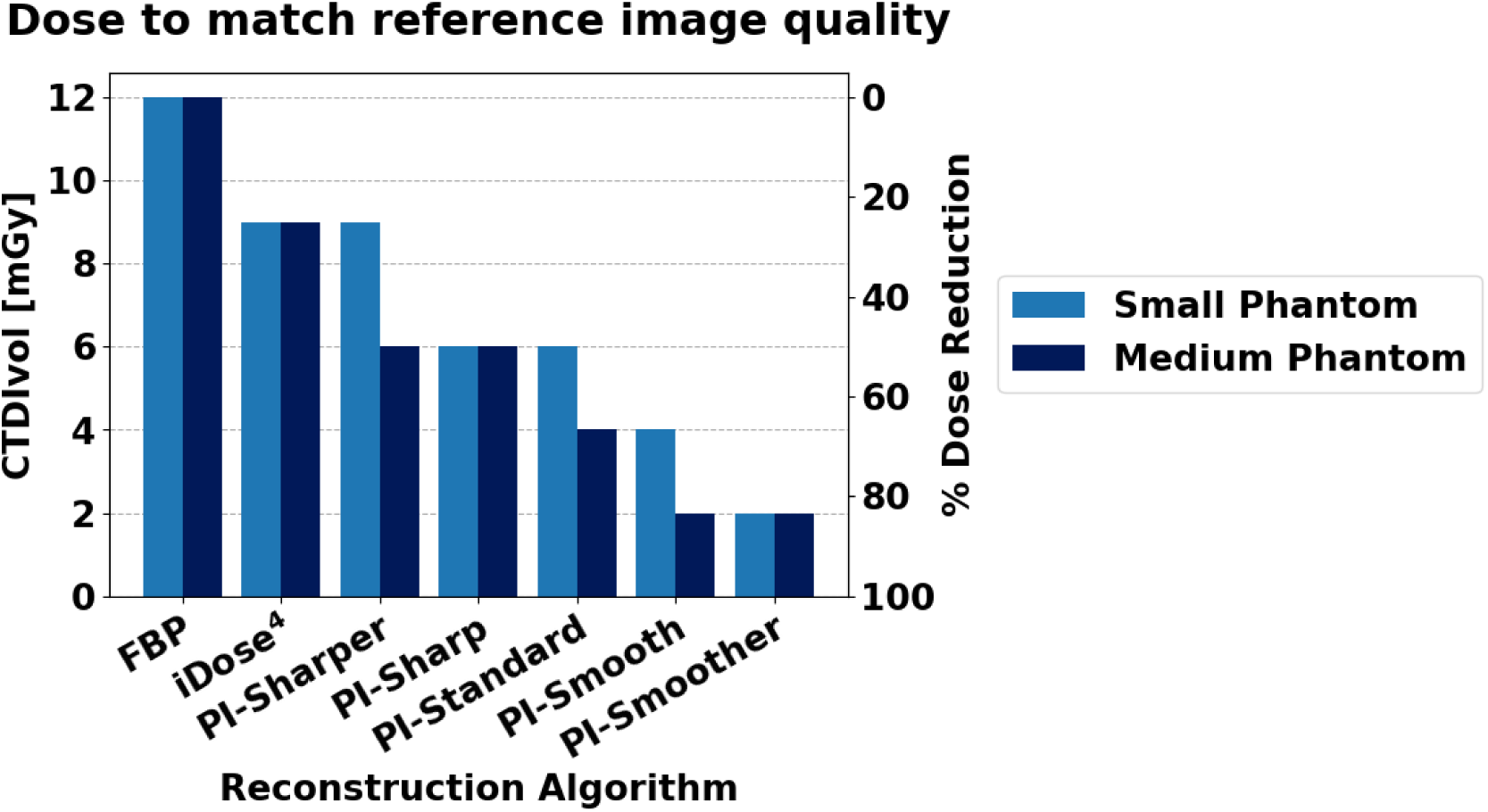
The minimum doses (left axis) required to match or exceed all image quality metrics of the reference images for each reconstruction algorithm, along with the corresponding percent dose reduction (right axis).

## 3 Discussion

This study examined the clinical imaging performance of a DLR algorithm, PI, compared to FBP and IR by utilizing a custom made patient derived PixelPrint lung phantom. The results show that PI is capable of dose reduction between 25-83% compared to FBP depending on the denoising level of the algorithm and phantom size. This suggests that in some cases PI can produce diagnostic level image quality even for CT scans acquired at lung cancer screening doses of < 3 mGy^35^. This could mean more effective lung cancer screening and/or reduced radiation burden.

The validity of using PixelPrint phantoms in the evaluation of DLR, and specifically PI, has also been demonstrated through this study. The results align with the current literature regarding image quality improvement, noise reduction, and dose reduction capabilities of DLR algorithms. A patient study by Greffier et. al^24^ looking at Precise Image for evaluating liver metastases^24^ demonstrated that the highest, most aggressive levels of PI (Smooth and Smoother) gave better scores for the lowest doses compared to the lower, less aggressive denoising levels of PI, the same trend found in this study. In a phantom study by Greffier et. al examining the use of PI in chest imaging, reported dose reductions were 58% for PI-Smooth and 83% for PI-Smoother compared to iDose^4^ Level 4^19^, again similar to the results of this study. Our study showed that in a medium sized phantom, if the results from MS SSIM are excluded, PI -Smooth has a 58% dose reduction potential and PI -Smoother an 88% dose reduction potential compared to iDose^4^ level 3. However, when MS SSIM is included the dose reduction potential becomes more conservative. Studies evaluating other DLR algorithms have also demonstrated reduced noise and improved lesion detectability in DLR compared to IR^18^ leading to a range of dose reduction estimates between 30-80%^1,20,21^. Overall, our results align closely with the results presented in previous studies evaluating DLR, which supports the validity of using PixelPrint phantoms for evaluation of DLR algorithms.

Furthermore, the results of this study demonstrate that PixelPrint phantoms provide additional information compared to patients and standard phantoms. Compared to patient studies, the use of PixelPrint phantoms allows for comparison of images over a wide range of doses beyond the dose range acceptable in clinical practice. This allows for the acquisition of ground truth data using a higher than standard radiation dose as well as repeated acquisition of increasingly lower dose data to probe dose limits more precisely. Additionally, there is no motion between acquisitions, which facilitates comparison between images using similarity metrics such as RMSE. Compared to traditional CT phantoms, the presence of clinically relevant structures and details in PixelPrint phantoms is advantageous because it enables comparison of reconstructed tissue structures via metrics like SSIM. Including these image similarity metrics resulted in more conservative dose reduction estimates compared to analysis including only general image quality metrics such as noise and CNR. This may be because DLR algorithms are inherently non-linear in nature and thus their performance is dependent on the specific characteristics of the objects being reconstructed. As a result, analyses on non-clinical structures such as those found in traditional CT phantoms cannot adequately capture these algorithms’ diagnostic imaging performance. PixelPrint phantoms can also provide additional information about patient size dependency in DLR dose reduction. A previous patient study using general image quality metrics showed that iDose^4^ achieved higher dose reduction in smaller patients versus larger patients^41^. In the present study, only considering the noise and CNR measurements results in the same trend while the inclusion of image similarity metrics results in a reversal of the trend such that the small phantom size has slightly reduced dose reduction potential. Finally, it has been reported that a possible concern with DLR is that if certain lesions are not well represented in training sets, these lesions may not be reconstructed accurately in DLR images^6^. This is not something that can be tested with standard geometric phantoms but can be easily investigated using PixelPrint phantoms with various lesions and known ground truth images. These findings suggest that the use of PixelPrint phantoms in conjunction with image similarity metrics provides valuable information which is not available from standard phantoms or patient studies alone for determining dose reduction capability.

The present study has a few limitations. First, PI was compared to only one denoising level of iDose^4^, the default level for lung imaging. To form a more robust understanding of the improvement that PI affords over IR, it would be valuable to compare PI to more denoising levels of iDose^4^. Second, this study only utilized one phantom and thus only one example of patient anatomy. Future studies involving more phantoms from different patients could improve our insights into the behavior of PI in different clinical scenarios and disease states. This could be especially useful in rarer or more unique clinical cases where patient data is limited. Third, this study only focuses on the performance of lung imaging since this PixelPrint phantom only included lung structures. Fourth, this study does not include a reader study involving subjective image quality scores. However, there are many existing studies that include a reader study portion and those results show good alignment with the results of the present study. Finally, the raw projection data corresponding to the patient images used to create the PixelPrint phantom was unavailable, preventing a direct comparison between PI performance on phantom data and its performance on the source patient data.

## 4 Conclusion

This study demonstrates the dose reduction capabilities of a DLR algorithm, Precise Image. PI has the capability of producing diagnostic image quality at up to 83% lower radiation dose, even surpassing the dose reduction capabilities of iterative reconstruction. These results are consistent with existing literature evaluating DLR. Images reconstructed using PI demonstrate not only improved noise and contrast compared to FBP and iterative reconstruction, but also improved structural accuracy of lung features such as GGO lesions. The use of PI can improve the clinical utility and viability of lower dose CT scans, ultimately improving patient care while reducing radiation exposure.

The PixelPrint phantom used in this study offers an improved testing environment with more realistic tissue structures and attenuation profiles compared to other CT phantoms. This is particularly important for the evaluation of non-linear reconstruction algorithms such as DLR. Thus, PixelPrint phantoms can elevate the clinical relevance of phantom evaluations of new and emerging CT technologies, which will lead to more rapid translation of these technologies into medical practice.

## Data Availability

All data produced in the present study are available upon reasonable request to the authors.

## 5 Acknowledgements

This work was partially funded by the National Institutes of Health through the following grants: R01EB030494, R01EB031592, R01HL166236, & R01CA249538. PN has received a hardware grant and research grant funding from Philips Healthcare. SH, AP, and EW are employees of Philips Healthcare. The other authors have no relevant conflicts of interest to disclose.

## Author contributions

Conception: JI, SH, PN

Design: JI, SH, KM, AP, LR, PN

Data acquisition: SH, EW Analysis: JI

Drafting: JI

Revising: JI, SH, KM, AP, OS, LL, GG, PN

## 7 Appendix

**Figure A1.**
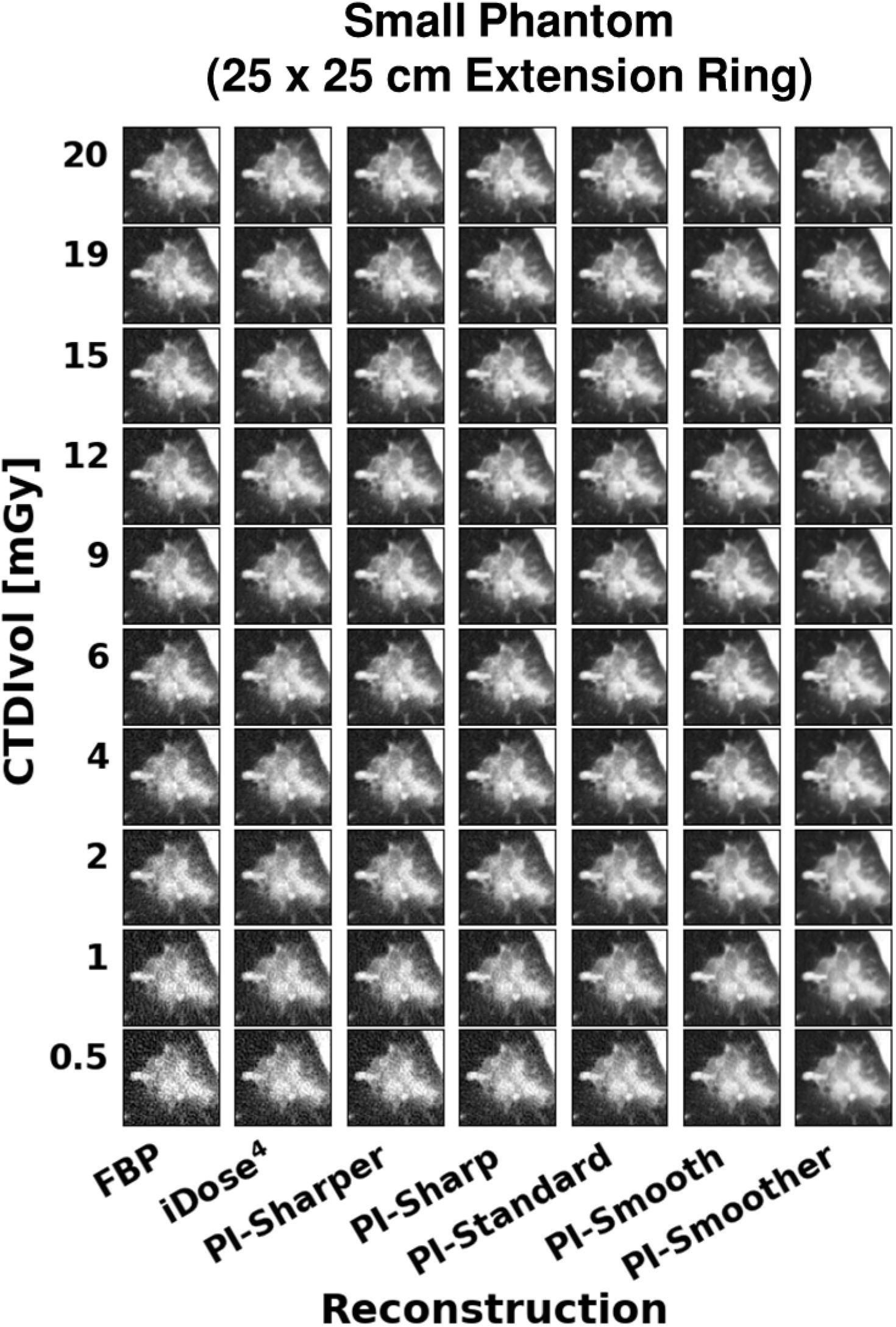
Images of the GGO lesion taken from the small phantom at each dose and reconstruction combination. WL: −500, WW: 1000.

**Figure A2.**
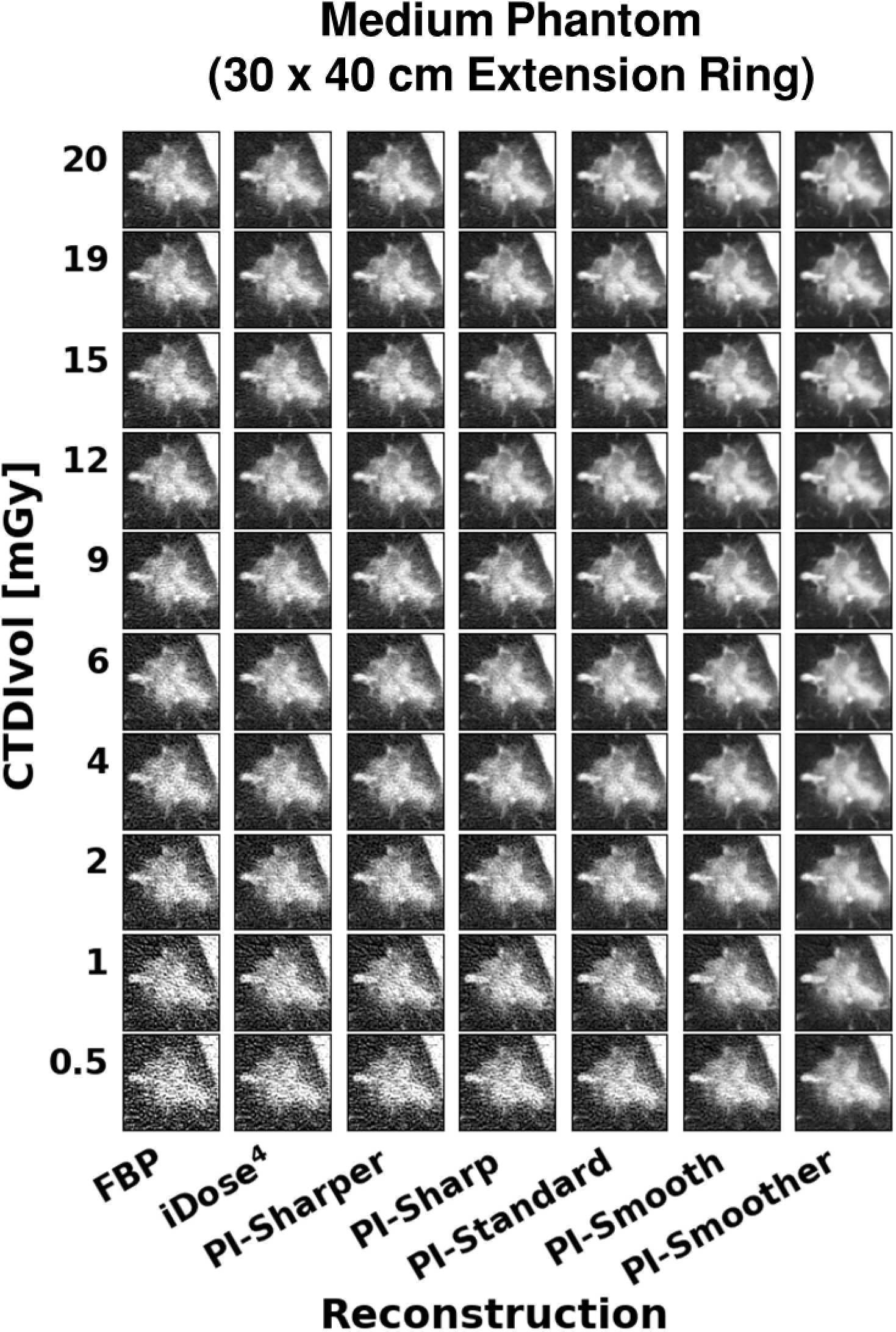
Images of the GGO lesion taken from the medium phantom at each dose and reconstruction combination. WL: −500, WW: 1000.

**Table A1.** Noise - Small Phantom. *t*-values and *p*-values obtained from the two-sample, one-tailed student’s t-test for each image metric calculated. The cells highlighted with red indicate where the *t*-value signifies worse performance (*t* < 0 for Noise and RMSE, *t* > 0 for CNR, SSIM, and MS SSIM) than the reference (FBP, 12 mGy). The cells highlighted in blue indicate where the *p*-value suggests a statistically significant result (*p* < 0.01). Cells above the thick black line have performance that is better than or not statistically different than the reference (*t* > 0 and/or *p* > 0.01). Cells below the thick black line have performance that is statistically worse than the reference (t < 0 and *p* < 0.01).

| CTDIvol [mGy] | FBP |  | iDose4, L3 |  | PI-Sharper |  | PI-Sharp |  | PI-Standard |  | PI-Smooth |  | PI-Smoother |  |
| --- | --- | --- | --- | --- | --- | --- | --- | --- | --- | --- | --- | --- | --- | --- |
|  | t | p | t | p | t | p | t | p | t | p | t | p | t | p |
| 19 | 6.761 | 0.00125 | 21.284 | 1.4E-05 | 18.823 | 2.3E-05 | 22.722 | 1.1E-05 | 26.661 | 5.9E-06 | 30.917 | 3.3E-06 | 34.427 | 2.1E-06 |
| 15 | 2.747 | 0.02578 | 17.985 | 2.8E-05 | 17.541 | 3.1E-05 | 22.060 | 1.2E-05 | 26.474 | 6E-06 | 31.028 | 3.2E-06 | 34.276 | 2.2E-06 |
| 12 | 0.000 | 0.5 | 14.665 | 6.3E-05 | 12.857 | 0.00011 | 17.298 | 3.3E-05 | 22.228 | 1.2E-05 | 27.898 | 4.9E-06 | 32.885 | 2.5E-06 |
| 9 | -3.864 | 0.00904 | 7.859 | 0.00071 | 7.521 | 0.00084 | 11.495 | 0.00016 | 15.927 | 4.5E-05 | 21.479 | 1.4E-05 | 27.262 | 5.4E-06 |
| 6 | -17.203 | 3.3E-05 | 3.561 | 0.01179 | 5.257 | 0.00313 | 12.392 | 0.00012 | 19.860 | 1.9E-05 | 28.348 | 4.6E-06 | 35.938 | 1.8E-06 |
| 4 | -9.091 | 0.00041 | -2.566 | 0.03114 | -1.426 | 0.1135 | 1.848 | 0.06919 | 5.911 | 0.00205 | 11.943 | 0.00014 | 19.397 | 2.1E-05 |
| 2 | -30.202 | 3.6E-06 | -18.046 | 2.8E-05 | -14.640 | 6.3E-05 | -7.544 | 0.00083 | 1.991 | 0.05866 | 16.937 | 3.6E-05 | 33.697 | 2.3E-06 |
| 1 | -131.203 | 1E-08 | -80.192 | 7.2E-08 | -63.965 | 1.8E-07 | -43.634 | 8.2E-07 | -24.505 | 8.2E-06 | -2.304 | 0.04129 | 16.855 | 3.6E-05 |
| 0.5 | -32.942 | 2.5E-06 | -27.526 | 5.2E-06 | -24.341 | 8.5E-06 | -20.110 | 1.8E-05 | -14.580 | 6.4E-05 | -5.465 | 0.00273 | 10.483 | 0.00023 |

**Table A2.**
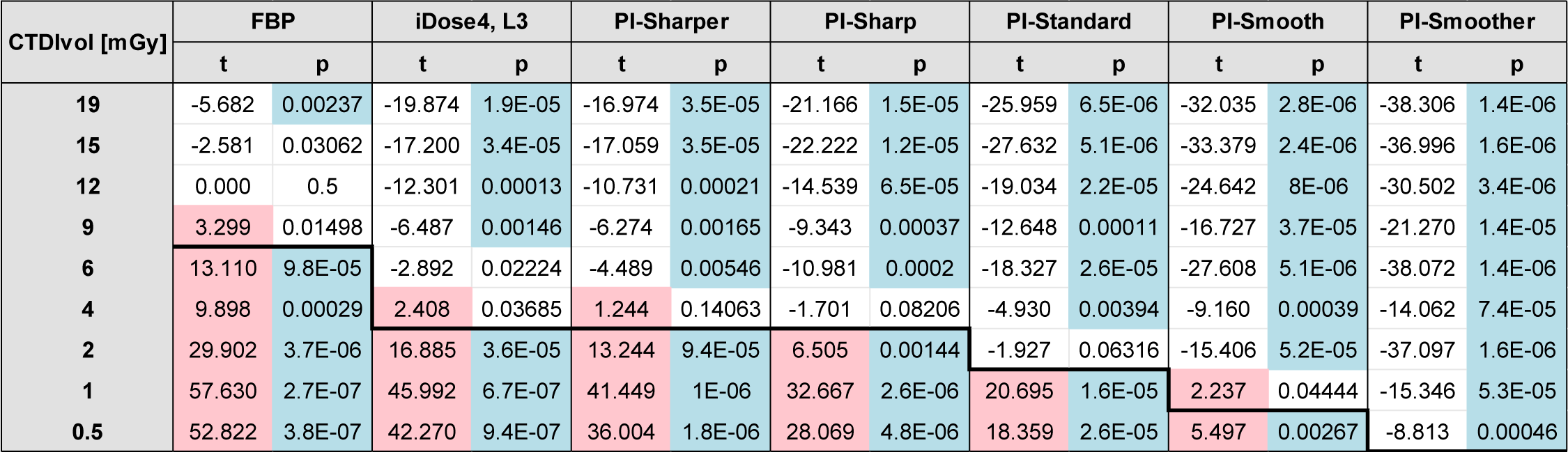
CNR - Small Phantom. *t*-values and *p*-values obtained from the two-sample, one-tailed student’s t-test for each image metric calculated. The cells highlighted with red indicate where the *t*-value signifies worse performance (*t* < 0 for Noise and RMSE, *t* > 0 for CNR, SSIM, and MS SSIM) than the reference (FBP, 12 mGy). The cells highlighted in blue indicate where the *p*-value suggests a statistically significant result (*p* < 0.01). Cells above the thick black line have performance that is better than or not statistically different than the reference (*t* > 0 and/or *p* > 0.01). Cells below the thick black line have performance that is statistically worse than the reference (t < 0 and *p* < 0.01).

**Table A3.** RMSE - Small Phantom. *t*-values and *p*-values obtained from the two-sample, one-tailed student’s t-test for each image metric calculated. The cells highlighted with red indicate where the *t*-value signifies worse performance (*t* < 0 for Noise and RMSE, *t* > 0 for CNR, SSIM, and MS SSIM) than the reference (FBP, 12 mGy). The cells highlighted in blue indicate where the *p*-value suggests a statistically significant result (*p* < 0.01). Cells above the thick black line have performance that is better than or not statistically different than the reference (*t* > 0 and/or *p* > 0.01). Cells below the thick black line have performance that is statistically worse than the reference (t < 0 and *p* < 0.01).

| CTDIvol [mGy] | FBP |  | iDose4, L3 |  | PI-Sharper |  | PI-Sharp |  | PI-Standard |  | PI-Smooth |  | PI-Smoother |  |
| --- | --- | --- | --- | --- | --- | --- | --- | --- | --- | --- | --- | --- | --- | --- |
|  | t | p | t | p | t | p | t | p | t | p | t | p | t | p |
| 19 | 9.022 | 0.00042 | 13.742 | 8.1E-05 | 15.565 | 5E-05 | 17.113 | 3.4E-05 | 18.411 | 2.6E-05 | 19.410 | 2.1E-05 | 19.635 | 2E-05 |
| 15 | 3.748 | 0.00999 | 9.611 | 0.00033 | 11.838 | 0.00015 | 13.908 | 7.7E-05 | 15.724 | 4.8E-05 | 17.299 | 3.3E-05 | 17.997 | 2.8E-05 |
| 12 | 0.000 | 0.5 | 8.267 | 0.00058 | 11.251 | 0.00018 | 14.048 | 7.4E-05 | 16.467 | 4E-05 | 18.533 | 2.5E-05 | 19.479 | 2E-05 |
| 9 | -10.673 | 0.00022 | 2.739 | 0.02598 | 8.158 | 0.00061 | 13.614 | 8.4E-05 | 18.675 | 2.4E-05 | 23.317 | 1E-05 | 25.617 | 6.9E-06 |
| 6 | -28.001 | 4.8E-06 | -10.363 | 0.00024 | -3.282 | 0.01522 | 3.641 | 0.01097 | 10.049 | 0.00028 | 16.124 | 4.3E-05 | 19.747 | 1.9E-05 |
| 4 | -57.470 | 2.7E-07 | -29.466 | 3.9E-06 | -19.627 | 2E-05 | -9.225 | 0.00038 | 0.162 | 0.43974 | 8.718 | 0.00048 | 13.679 | 8.3E-05 |
| 2 | -117.216 | 1.6E-08 | -73.213 | 1E-07 | -58.057 | 2.6E-07 | -39.635 | 1.2E-06 | -21.997 | 1.3E-05 | -4.917 | 0.00397 | 5.723 | 0.00231 |
| 1 | -207.496 | 1.6E-09 | -146.938 | 6.4E-09 | -122.796 | 1.3E-08 | -94.716 | 3.7E-08 | -65.543 | 1.6E-07 | -32.787 | 2.6E-06 | -6.935 | 0.00113 |
| 0.5 | -189.715 | 2.3E-09 | -153.389 | 5.4E-09 | -134.644 | 9.1E-09 | -113.199 | 1.8E-08 | -88.169 | 5E-08 | -55.673 | 3.1E-07 | -23.898 | 9.1E-06 |

**Table A4.** SSIM - Small Phantom. *t*-values and *p*-values obtained from the two-sample, one-tailed student’s t-test for each image metric calculated. The cells highlighted with red indicate where the *t*-value signifies worse performance (*t* < 0 for Noise and RMSE, *t* > 0 for CNR, SSIM, and MS SSIM) than the reference (FBP, 12 mGy). The cells highlighted in blue indicate where the *p*-value suggests a statistically significant result (*p* < 0.01). Cells above the thick black line have performance that is better than or not statistically different than the reference (*t* > 0 and/or *p* > 0.01). Cells below the thick black line have performance that is statistically worse than the reference (t < 0 and *p* < 0.01).

| CTDIvol [mGy] | FBP |  | iDose4, L3 |  | PI-Sharper |  | PI-Sharp |  | PI-Standard |  | PI-Smooth |  | PI-Smoother |  |
| --- | --- | --- | --- | --- | --- | --- | --- | --- | --- | --- | --- | --- | --- | --- |
|  | t | p | t | p | t | p | t | p | t | p | t | p | t | p |
| 19 | -6.253 | 0.00167 | -9.816 | 0.0003 | -11.165 | 0.00018 | -12.508 | 0.00012 | -13.722 | 8.2E-05 | -14.823 | 6E-05 | -15.470 | 5.1E-05 |
| 15 | -3.075 | 0.01855 | -8.389 | 0.00055 | -9.975 | 0.00028 | -11.839 | 0.00015 | -13.239 | 9.4E-05 | -14.524 | 6.5E-05 | -14.784 | 6.1E-05 |
| 12 | 0.000 | 0.5 | -6.636 | 0.00134 | -8.185 | 0.00061 | -10.298 | 0.00025 | -12.083 | 0.00013 | -13.797 | 8E-05 | -14.620 | 6.4E-05 |
| 9 | 8.766 | 0.00047 | -0.843 | 0.22325 | -3.990 | 0.00813 | -7.639 | 0.00079 | -10.956 | 0.0002 | -14.058 | 7.4E-05 | -15.896 | 4.6E-05 |
| 6 | 15.212 | 5.4E-05 | 5.687 | 0.00236 | 2.790 | 0.02465 | -1.337 | 0.12611 | -5.874 | 0.0021 | -10.849 | 0.0002 | -13.903 | 7.8E-05 |
| 4 | 23.776 | 9.3E-06 | 13.918 | 7.7E-05 | 9.657 | 0.00032 | 5.562 | 0.00256 | 1.019 | 0.18283 | -4.643 | 0.00486 | -8.888 | 0.00044 |
| 2 | 50.550 | 4.6E-07 | 27.778 | 5E-06 | 32.064 | 2.8E-06 | 24.165 | 8.7E-06 | 14.866 | 6E-05 | 4.218 | 0.00675 | -3.582 | 0.01156 |
| 1 | 76.733 | 8.6E-08 | 47.351 | 5.9E-07 | 63.213 | 1.9E-07 | 56.416 | 3E-07 | 46.731 | 6.3E-07 | 28.522 | 4.5E-06 | 6.650 | 0.00133 |
| 0.5 | 157.740 | 4.8E-09 | 122.256 | 1.3E-08 | 141.228 | 7.5E-09 | 125.567 | 1.2E-08 | 97.477 | 3.3E-08 | 48.559 | 5.4E-07 | 14.264 | 7E-05 |

**Table A5.** MS SSIM - Small Phantom. *t*-values and *p*-values obtained from the two-sample, one-tailed student’s t-test for each image metric calculated. The cells highlighted with red indicate where the *t*-value signifies worse performance (*t* < 0 for Noise and RMSE, *t* > 0 for CNR, SSIM, and MS SSIM) than the reference (FBP, 12 mGy). The cells highlighted in blue indicate where the *p*-value suggests a statistically significant result (*p* < 0.01). Cells above the thick black line have performance that is better than or not statistically different than the reference (*t* > 0 and/or *p* > 0.01). Cells below the thick black line have performance that is statistically worse than the reference (t < 0 and *p* < 0.01).

| CTDIvol [mGy] | FBP |  | iDose4, L3 |  | PI-Sharper |  | PI-Sharp |  | PI-Standard |  | PI-Smooth |  | PI-Smoother |  |
| --- | --- | --- | --- | --- | --- | --- | --- | --- | --- | --- | --- | --- | --- | --- |
|  | t | p | t | p | t | p | t | p | t | p | t | p | t | p |
| 19 | -8.576 | 0.00051 | -13.895 | 7.8E-05 | -15.959 | 4.5E-05 | -18.335 | 2.6E-05 | -20.725 | 1.6E-05 | -23.408 | 9.9E-06 | -26.204 | 6.3E-06 |
| 15 | -4.464 | 0.00556 | -12.587 | 0.00011 | -14.074 | 7.4E-05 | -16.237 | 4.2E-05 | -17.334 | 3.3E-05 | -18.071 | 2.8E-05 | -17.353 | 3.2E-05 |
| 12 | 0.000 | 0.5 | -10.559 | 0.00023 | -12.470 | 0.00012 | -15.250 | 5.4E-05 | -17.268 | 3.3E-05 | -19.559 | 2E-05 | -20.828 | 1.6E-05 |
| 9 | 11.911 | 0.00014 | -2.052 | 0.0547 | -5.914 | 0.00205 | -10.778 | 0.00021 | -14.870 | 6E-05 | -18.378 | 2.6E-05 | -20.857 | 1.6E-05 |
| 6 | 28.021 | 4.8E-06 | 9.674 | 0.00032 | 6.062 | 0.00187 | -0.552 | 0.30515 | -7.552 | 0.00082 | -14.267 | 7E-05 | -16.820 | 3.7E-05 |
| 4 | 51.394 | 4.3E-07 | 27.620 | 5.1E-06 | 21.278 | 1.4E-05 | 14.183 | 7.2E-05 | 5.828 | 0.00216 | -3.753 | 0.00995 | -8.802 | 0.00046 |
| 2 | 57.907 | 2.7E-07 | 30.306 | 3.5E-06 | 60.686 | 2.2E-07 | 53.371 | 3.7E-07 | 34.361 | 2.1E-06 | 18.110 | 2.7E-05 | 2.431 | 0.03595 |
| 1 | 48.771 | 5.3E-07 | 30.914 | 3.3E-06 | 53.438 | 3.7E-07 | 55.382 | 3.2E-07 | 57.712 | 2.7E-07 | 52.878 | 3.8E-07 | 19.623 | 2E-05 |
| 0.5 | 167.543 | 3.8E-09 | 124.102 | 1.3E-08 | 243.779 | 8.5E-10 | 210.586 | 1.5E-09 | 152.992 | 5.5E-09 | 71.409 | 1.2E-07 | 29.596 | 3.9E-06 |

**Table A6.**
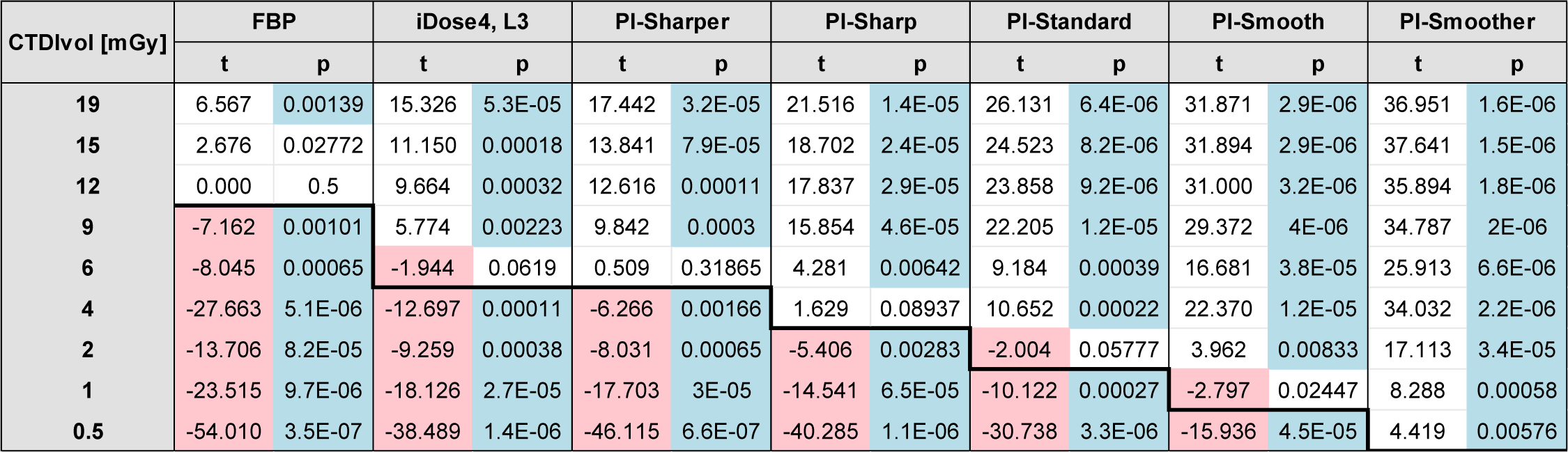
Noise - Medium Phantom. *t*-values and *p*-values obtained from the two-sample, one-tailed student’s t-test for each image metric calculated. The cells highlighted with red indicate where the *t*-value signifies worse performance (*t* < 0 for Noise and RMSE, *t* > 0 for CNR, SSIM, and MS SSIM) than the reference (FBP, 12 mGy). The cells highlighted in blue indicate where the *p*-value suggests a statistically significant result (*p* < 0.01). Cells above the thick black line have performance that is better than or not statistically different than the reference (*t* > 0 and/or *p* > 0.01). Cells below the thick black line have performance that is statistically worse than the reference (t < 0 and *p* < 0.01).

**Table A7.** CNR - Medium Phantom. *t*-values and *p*-values obtained from the two-sample, one-tailed student’s t-test for each image metric calculated. The cells highlighted with red indicate where the *t*-value signifies worse performance (*t* < 0 for Noise and RMSE, *t* > 0 for CNR, SSIM, and MS SSIM) than the reference (FBP, 12 mGy). The cells highlighted in blue indicate where the *p*-value suggests a statistically significant result (*p* < 0.01). Cells above the thick black line have performance that is better than or not statistically different than the reference (*t* > 0 and/or *p* > 0.01). Cells below the thick black line have performance that is statistically worse than the reference (t < 0 and *p* < 0.01).

| CTDIvol [mGy] | FBP |  | iDose4, L3 |  | PI-Sharper |  | PI-Sharp |  | PI-Standard |  | PI-Smooth |  | PI-Smoother |  |
| --- | --- | --- | --- | --- | --- | --- | --- | --- | --- | --- | --- | --- | --- | --- |
|  | t | p | t | p | t | p | t | p | t | p | t | p | t | p |
| 19 | -6.450 | 0.00149 | -14.883 | 5.9E-05 | -17.073 | 3.5E-05 | -21.357 | 1.4E-05 | -27.082 | 5.5E-06 | -37.861 | 1.5E-06 | -61.046 | 2.2E-07 |
| 15 | -2.717 | 0.02657 | -10.675 | 0.00022 | -13.389 | 9E-05 | -18.527 | 2.5E-05 | -26.468 | 6.1E-06 | -46.331 | 6.5E-07 | -89.628 | 4.6E-08 |
| 12 | 0.000 | 0.5 | -9.241 | 0.00038 | -12.397 | 0.00012 | -18.232 | 2.7E-05 | -26.665 | 5.9E-06 | -41.654 | 9.9E-07 | -44.483 | 7.6E-07 |
| 9 | 6.087 | 0.00184 | -6.176 | 0.00175 | -11.436 | 0.00017 | -20.486 | 1.7E-05 | -32.346 | 2.7E-06 | -44.840 | 7.4E-07 | -39.780 | 1.2E-06 |
| 6 | 9.095 | 0.00041 | 1.965 | 0.0604 | -0.695 | 0.26264 | -4.191 | 0.0069 | -7.926 | 0.00069 | -12.200 | 0.00013 | -16.082 | 4.4E-05 |
| 4 | 18.762 | 2.4E-05 | 10.146 | 0.00027 | 5.223 | 0.00321 | -1.669 | 0.08518 | -10.902 | 0.0002 | -24.912 | 7.7E-06 | -42.184 | 9.4E-07 |
| 2 | 19.823 | 1.9E-05 | 13.115 | 9.8E-05 | 10.515 | 0.00023 | 6.561 | 0.0014 | 2.064 | 0.05398 | -3.586 | 0.01152 | -10.958 | 0.0002 |
| 1 | 32.666 | 2.6E-06 | 26.655 | 5.9E-06 | 23.702 | 9.4E-06 | 19.166 | 2.2E-05 | 12.859 | 0.00011 | 3.042 | 0.01916 | -6.220 | 0.0017 |
| 0.5 | 41.139 | 1E-06 | 36.777 | 1.6E-06 | 34.589 | 2.1E-06 | 31.148 | 3.2E-06 | 25.849 | 6.7E-06 | 15.081 | 5.6E-05 | -4.178 | 0.00697 |

**Table A8.** RMSE - Medium Phantom. *t*-values and *p*-values obtained from the two-sample, one-tailed student’s t-test for each image metric calculated. The cells highlighted with red indicate where the *t*-value signifies worse performance (*t* < 0 for Noise and RMSE, *t* > 0 for CNR, SSIM, and MS SSIM) than the reference (FBP, 12 mGy). The cells highlighted in blue indicate where the *p*-value suggests a statistically significant result (*p* < 0.01). Cells above the thick black line have performance that is better than or not statistically different than the reference (*t* > 0 and/or *p* > 0.01). Cells below the thick black line have performance that is statistically worse than the reference (t < 0 and *p* < 0.01).

| CTDIvol [mGy] | FBP |  | iDose4, L3 |  | PI-Sharper |  | PI-Sharp |  | PI-Standard |  | PI-Smooth |  | PI-Smoother |  |
| --- | --- | --- | --- | --- | --- | --- | --- | --- | --- | --- | --- | --- | --- | --- |
|  | t | p | t | p | t | p | t | p | t | p | t | p | t | p |
| 19 | 44.258 | 7.8E-07 | 73.694 | 1E-07 | 86.812 | 5.3E-08 | 100.083 | 3E-08 | 113.486 | 1.8E-08 | 128.633 | 1.1E-08 | 142.164 | 7.3E-09 |
| 15 | 21.459 | 1.4E-05 | 61.105 | 2.1E-07 | 79.889 | 7.4E-08 | 100.645 | 2.9E-08 | 121.589 | 1.4E-08 | 143.956 | 7E-09 | 161.626 | 4.4E-09 |
| 12 | 0.000 | 0.5 | 53.509 | 3.7E-07 | 74.716 | 9.6E-08 | 97.681 | 3.3E-08 | 120.655 | 1.4E-08 | 146.253 | 6.6E-09 | 167.884 | 3.8E-09 |
| 9 | -49.981 | 4.8E-07 | 25.468 | 7.1E-06 | 51.251 | 4.3E-07 | 77.265 | 8.4E-08 | 101.083 | 2.9E-08 | 125.004 | 1.2E-08 | 144.281 | 6.9E-09 |
| 6 | -74.652 | 9.6E-08 | -26.617 | 5.9E-06 | -0.645 | 0.27694 | 31.399 | 3.1E-06 | 68.483 | 1.4E-07 | 112.823 | 1.9E-08 | 148.735 | 6.1E-09 |
| 4 | -280.873 | 4.8E-10 | -156.561 | 5E-09 | -87.198 | 5.2E-08 | -25.306 | 7.2E-06 | 36.939 | 1.6E-06 | 105.108 | 2.5E-08 | 159.904 | 4.6E-09 |
| 2 | -166.211 | 3.9E-09 | -127.538 | 1.1E-08 | -114.549 | 1.7E-08 | -88.419 | 4.9E-08 | -48.958 | 5.2E-07 | 23.809 | 9.2E-06 | 110.499 | 2E-08 |
| 1 | -383.558 | 1.4E-10 | -312.656 | 3.1E-10 | -283.320 | 4.7E-10 | -235.633 | 9.7E-10 | -173.495 | 3.3E-09 | -74.744 | 9.6E-08 | 62.681 | 1.9E-07 |
| 0.5 | -218.662 | 1.3E-09 | -181.638 | 2.8E-09 | -176.841 | 3.1E-09 | -156.027 | 5.1E-09 | -127.778 | 1.1E-08 | -82.302 | 6.5E-08 | -11.521 | 0.00016 |

**Table A9.** SSIM - Medium Phantom. *t*-values and *p*-values obtained from the two-sample, one-tailed student’s t-test for each image metric calculated. The cells highlighted with red indicate where the *t*-value signifies worse performance (*t* < 0 for Noise and RMSE, *t* > 0 for CNR, SSIM, and MS SSIM) than the reference (FBP, 12 mGy). The cells highlighted in blue indicate where the *p*-value suggests a statistically significant result (*p* < 0.01). Cells above the thick black line have performance that is better than or not statistically different than the reference (*t* > 0 and/or *p* > 0.01). Cells below the thick black line have performance that is statistically worse than the reference (t < 0 and *p* < 0.01).

| CTDIvol [mGy] | FBP |  | iDose4, L3 |  | PI-Sharper |  | PI-Sharp |  | PI-Standard |  | PI-Smooth |  | PI-Smoother |  |
| --- | --- | --- | --- | --- | --- | --- | --- | --- | --- | --- | --- | --- | --- | --- |
|  | t | p | t | p | t | p | t | p | t | p | t | p | t | p |
| 19 | -19.370 | 2.1E-05 | -31.832 | 2.9E-06 | -38.489 | 1.4E-06 | -48.070 | 5.6E-07 | -51.121 | 4.4E-07 | -52.955 | 3.8E-07 | -56.114 | 3E-07 |
| 15 | -7.318 | 0.00093 | -23.726 | 9.4E-06 | -29.889 | 3.7E-06 | -38.887 | 1.3E-06 | -42.319 | 9.3E-07 | -51.829 | 4.1E-07 | -64.686 | 1.7E-07 |
| 12 | 0.000 | 0.5 | -27.447 | 5.2E-06 | -40.491 | 1.1E-06 | -57.749 | 2.7E-07 | -70.720 | 1.2E-07 | -61.592 | 2.1E-07 | -73.123 | 1E-07 |
| 9 | 15.645 | 4.9E-05 | -5.426 | 0.0028 | -16.187 | 4.3E-05 | -30.130 | 3.6E-06 | -43.755 | 8.2E-07 | -57.682 | 2.7E-07 | -64.132 | 1.8E-07 |
| 6 | 14.748 | 6.2E-05 | 4.581 | 0.00509 | 1.407 | 0.11611 | -4.881 | 0.00408 | -14.256 | 7E-05 | -33.151 | 2.5E-06 | -48.538 | 5.4E-07 |
| 4 | 68.332 | 1.4E-07 | 34.075 | 2.2E-06 | 28.008 | 4.8E-06 | 11.720 | 0.00015 | -7.595 | 0.00081 | -35.739 | 1.8E-06 | -62.100 | 2E-07 |
| 2 | 48.862 | 5.2E-07 | 33.319 | 2.4E-06 | 33.102 | 2.5E-06 | 25.422 | 7.1E-06 | 15.173 | 5.5E-05 | -2.201 | 0.04628 | -31.278 | 3.1E-06 |
| 1 | 206.702 | 1.6E-09 | 139.426 | 7.9E-09 | 152.245 | 5.6E-09 | 125.979 | 1.2E-08 | 93.240 | 4E-08 | 41.728 | 9.9E-07 | -10.140 | 0.00027 |
| 0.5 | 232.325 | 1E-09 | 153.242 | 5.4E-09 | 246.323 | 8.1E-10 | 245.382 | 8.3E-10 | 163.871 | 4.2E-09 | 84.515 | 5.9E-08 | 17.028 | 3.5E-05 |

**Table A10.** MS SSIM - Medium Phantom. *t*-values and *p*-values obtained from the two-sample, one-tailed student’s t-test for each image metric calculated. The cells highlighted with red indicate where the *t*-value signifies worse performance (*t* < 0 for Noise and RMSE, *t* > 0 for CNR, SSIM, and MS SSIM) than the reference (FBP, 12 mGy). The cells highlighted in blue indicate where the *p*-value suggests a statistically significant result (*p* < 0.01). Cells above the thick black line have performance that is better than or not statistically different than the reference (*t* > 0 and/or *p* > 0.01). Cells below the thick black line have performance that is statistically worse than the reference (t < 0 and *p* < 0.01).

| CTDIvol [mGy] | FBP |  | iDose4, L3 |  | PI-Sharper |  | PI-Sharp |  | PI-Standard |  | PI-Smooth |  | PI-Smoother |  |
| --- | --- | --- | --- | --- | --- | --- | --- | --- | --- | --- | --- | --- | --- | --- |
|  | t | p | t | p | t | p | t | p | t | p | t | p | t | p |
| 19 | -16.977 | 3.5E-05 | -27.404 | 5.3E-06 | -30.150 | 3.6E-06 | -36.641 | 1.7E-06 | -39.635 | 1.2E-06 | -40.705 | 1.1E-06 | -40.808 | 1.1E-06 |
| 15 | -8.549 | 0.00051 | -22.780 | 1.1E-05 | -29.635 | 3.9E-06 | -35.996 | 1.8E-06 | -36.974 | 1.6E-06 | -40.680 | 1.1E-06 | -42.979 | 8.8E-07 |
| 12 | 0.000 | 0.5 | -13.394 | 9E-05 | -20.279 | 1.7E-05 | -28.750 | 4.4E-06 | -37.559 | 1.5E-06 | -36.619 | 1.7E-06 | -39.246 | 1.3E-06 |
| 9 | 8.833 | 0.00045 | -6.350 | 0.00158 | -11.583 | 0.00016 | -19.357 | 2.1E-05 | -27.832 | 5E-06 | -34.745 | 2E-06 | -39.366 | 1.2E-06 |
| 6 | 18.062 | 2.8E-05 | 3.675 | 0.01065 | 2.158 | 0.04857 | -5.776 | 0.00223 | -16.554 | 3.9E-05 | -28.946 | 4.2E-06 | -28.954 | 4.2E-06 |
| 4 | 55.655 | 3.1E-07 | 22.686 | 1.1E-05 | 26.618 | 5.9E-06 | 14.356 | 6.8E-05 | 0.110 | 0.45874 | -18.159 | 2.7E-05 | -28.857 | 4.3E-06 |
| 2 | 16.350 | 4.1E-05 | 11.286 | 0.00018 | 13.231 | 9.4E-05 | 11.330 | 0.00017 | 8.466 | 0.00053 | 1.754 | 0.07716 | -12.498 | 0.00012 |
| 1 | 97.635 | 3.3E-08 | 77.421 | 8.3E-08 | 59.528 | 2.4E-07 | 47.219 | 6E-07 | 37.883 | 1.4E-06 | 31.359 | 3.1E-06 | 8.942 | 0.00043 |
| 0.5 | 106.521 | 2.3E-08 | 69.036 | 1.3E-07 | 94.496 | 3.8E-08 | 84.870 | 5.8E-08 | 86.032 | 5.5E-08 | 55.308 | 3.2E-07 | 26.770 | 5.8E-06 |

## Notes

### Competing Interest Statement

PN has received a hardware grant from Philips Healthcare. PN receives research grant funding from Philips Healthcare. SH, AP, and EW are employees of Philips Healthcare. The other authors have no relevant conflicts of interest to disclose.

### Funding Statement

We acknowledge support through the National Institutes of Health (R01EB030494, R01EB031592, R01HL166236, & R01CA249538).

### Author Declarations

The Institutional Review Board (IRB) at the University of Pennsylvania approved this retrospective study.

## References

1. Koetzier LR, Mastrodicasa D, Szczykutowicz TP, et al. Deep Learning Image Reconstruction for CT: Technical Principles and Clinical Prospects. Radiology. Mar 2023;306(3):e221257. doi:10.1148/radiol.221257

2. Willemink MJ, Noël PB. The evolution of image reconstruction for CT—from filtered back projection to artificial intelligence. European Radiology. 2019/05/01 2019;29(5):2185-2195. doi:10.1007/s00330-018-5810-7

3. Brenner DJ, Hall EJ. Computed Tomography — An Increasing Source of Radiation Exposure. New England Journal of Medicine. 2007;357(22):2277–2284. doi:10.1056/NEJMra072149

4. Miglioretti DL, Johnson E, Williams A, et al. The Use of Computed Tomography in Pediatrics and the Associated Radiation Exposure and Estimated Cancer Risk. JAMA Pediatrics. 2013;167(8):700–707. doi:10.1001/jamapediatrics.2013.311

5. Son W, Kim M, Hwang JY, et al. Comparison of a Deep Learning-Based Reconstruction Algorithm with Filtered Back Projection and Iterative Reconstruction Algorithms for Pediatric Abdominopelvic CT. Korean J Radiol. Jul 2022;23(7):752–762. doi:10.3348/kjr.2021.0466

6. Nagayama Y, Sakabe D, Goto M, et al. Deep Learning-based Reconstruction for Lower-Dose Pediatric CT: Technical Principles, Image Characteristics, and Clinical Implementations. Radiographics. Nov-Dec 2021;41(7):1936–1953. doi:10.1148/rg.2021210105

7. Sun J, Li H, Li J, et al. Improving the image quality of pediatric chest CT angiography with low radiation dose and contrast volume using deep learning image reconstruction. Quantitative Imaging in Medicine and Surgery. 2021;11(7):3051.

8. Willemink MJ, de Jong PA, Leiner T, et al. Iterative reconstruction techniques for computed tomography Part 1: Technical principles. European Radiology. 2013/06/01 2013;23(6):1623–1631. doi:10.1007/s00330-012-2765-y

9. Philips. AI for significantly lower dose and improved image quality. 2021;

10. Kang E, Min J, Ye JC. A deep convolutional neural network using directional wavelets for low-dose X-ray CT reconstruction. Medical physics. 2017;44(10):e360–e375.

11. Chen H, Zhang Y, Kalra MK, et al. Low-dose CT with a residual encoder-decoder convolutional neural network. IEEE transactions on medical imaging. 2017;36(12):2524–2535.

12. Wolterink JM, Leiner T, Viergever MA, Išgum I. Generative Adversarial Networks for Noise Reduction in Low-Dose CT. IEEE Transactions on Medical Imaging. 2017;36(12):2536–2545. doi:10.1109/TMI.2017.2708987

13. Wu D, Kim K, Fakhri GE, Li Q. Iterative Low-Dose CT Reconstruction With Priors Trained by Artificial Neural Network. IEEE Transactions on Medical Imaging. 2017;36(12):2479–2486. doi:10.1109/TMI.2017.2753138

14. Yang Q, Yan P, Zhang Y, et al. Low-Dose CT Image Denoising Using a Generative Adversarial Network With Wasserstein Distance and Perceptual Loss. IEEE Transactions on Medical Imaging. 2018;37(6):1348–1357. doi:10.1109/TMI.2018.2827462

15. Bao P, Xia W, Yang K, et al. Convolutional Sparse Coding for Compressed Sensing CT Reconstruction. IEEE Transactions on Medical Imaging. 2019;38(11):2607–2619. doi:10.1109/TMI.2019.2906853

16. Hsieh J, Liu E, Net B, Tang J, Thibault J-B, Sahney S. A new era of image reconstruction: TrueFidelity™. White Paper (JB68676XX), GE Healthcare. 2019;

17. Boedeker K. AiCE Deep Learning Reconstruction: Bringing the power of Ultra-High Resolution CT to routine imaging. Canon Medical Systems. 2021.

18. Nakamura Y, Higaki T, Tatsugami F, et al. Deep Learning-based CT Image Reconstruction: Initial Evaluation Targeting Hypovascular Hepatic Metastases. Radiol Artif Intell. Nov 2019;1(6):e180011. doi:10.1148/ryai.2019180011

19. Greffier J, Si-Mohamed S, Frandon J, et al. Impact of an artificial intelligence deep-learning reconstruction algorithm for CT on image quality and potential dose reduction: A phantom study. Med Phys. Aug 2022;49(8):5052–5063. doi:10.1002/mp.15807

20. Miyata T, Yanagawa M, Kikuchi N, et al. The evaluation of the reduction of radiation dose via deep learning-based reconstruction for cadaveric human lung CT images. Sci Rep. Jul 20 2022;12(1):12422. doi:10.1038/s41598-022-16798-9

21. Park J, Shin J, Min IK, Bae H, Kim YE, Chung YE. Image Quality and Lesion Detectability of Lower-Dose Abdominopelvic CT Obtained Using Deep Learning Image Reconstruction. Korean J Radiol. Apr 2022;23(4):402–412. doi:10.3348/kjr.2021.0683

22. Greffier J, Frandon J, Si-Mohamed S, et al. Comparison of two deep learning image reconstruction algorithms in chest CT images: A task-based image quality assessment on phantom data. Diagnostic and Interventional Imaging. 2022/01/01/ 2022;103(1):21–30. doi:htps://doi.org/10.1016/j.diii.2021.08.001

23. Mikayama R, Shirasaka T, Kojima T, et al. Deep-learning reconstruction for ultra-low-dose lung CT: volumetric measurement accuracy and reproducibility of artificial ground-glass nodules in a phantom study. The British Journal of Radiology. 2022;95(1130):20210915.

24. Greffier J, Durand Q, Serrand C, et al. First Results of a New Deep Learning Reconstruction Algorithm on Image Quality and Liver Metastasis Conspicuity for Abdominal Low-Dose CT. Diagnostics. 2023;13(6):1182.

25. Akagi M, Nakamura Y, Higaki T, et al. Deep learning reconstruction improves image quality of abdominal ultra-high-resolution CT. European Radiology. 2019/11/01 2019;29(11):6163–6171. doi:10.1007/s00330-019-06170-3

26. Li J, Wang W, Tivnan M, et al. Local linearity analysis of deep learning CT denoising algorithms. SPIE;

27. Lyu P, Li Z, Chen Y, et al. Deep learning reconstruction CT for liver metastases: low-dose dual-energy vs standard-dose single-energy. European Radiology. 2023/08/02 2023;doi:10.1007/s00330-023-10033-3

28. Infante M, Lutman RF, Imparato S, et al. Differential diagnosis and management of focal ground-glass opacities. European Respiratory Journal. 2009;33(4):821–827. doi:10.1183/09031936.00047908

29. Mei K, Geagan M, Shapira N, et al. PixelPrint: Three-dimensional printing of patient-specific soft tissue and bone phantoms for CT. Proc SPIE Int Soc Opt Eng. Jun 2022;12304doi:10.1117/12.2647008

30. Shapira N, Donovan K, Mei K, et al. Three-dimensional printing of patient-specific computed tomography lung phantoms: a reader study. PNAS Nexus. Mar 2023;2(3):pgad026. doi:10.1093/pnasnexus/pgad026

31. Shapira N, Donovan K, Mei K, et al. PixelPrint: Three-dimensional printing of realistic patient-specific lung phantoms for CT imaging. Proc SPIE Int Soc Opt Eng. Feb-Mar 2022;12031doi:10.1117/12.2611805

32. Mei K, Geagan M, Roshkovan L, et al. Three-dimensional printing of patient-specific lung phantoms for CT imaging: Emulating lung tissue with accurate atenuation profiles and textures. Medical Physics. 2022;49(2):825–835. doi:10.1002/mp.15407

33. Hofmanninger J, Prayer F, Pan J, Röhrich S, Prosch H, Langs G. Automatic lung segmentation in routine imaging is primarily a data diversity problem, not a methodology problem. European Radiology Experimental. 2020-12-01 2020;4(1)doi:10.1186/s41747-020-00173-2

34. Radiology ACo. ACR–AAPM–SPR PRACTICE PARAMETER FOR DIAGNOSTIC REFERENCE LEVELS AND ACHIEVABLE DOSES IN MEDICAL X-RAY IMAGING. 2018.

35. Kazerooni EA, Austin JH, Black WC, et al. ACR-STR practice parameter for the performance and reporting of lung cancer screening thoracic computed tomography (CT): 2014 (Resolution 4). J Thorac Imaging. Sep 2014;29(5):310–6. doi:10.1097/RTI.0000000000000097

36. Wang Z, Bovik AC, Sheikh HR, Simoncelli EP. Image quality assessment: from error visibility to structural similarity. IEEE Trans Image Process. Apr 2004;13(4):600–12. doi:10.1109/tip.2003.819861

37. Wang Z, Simoncelli EP, Bovik AC. Multiscale structural similarity for image quality assessment. 2003:1398–1402 Vol.2.

38. van der Walt S, Schonberger JL, Nunez-Iglesias J, et al. scikit-image: image processing in Python. Peerj. Jun 19 2014;2doi:ARTN e453 10.7717/peerj.453

39. Pessoa J. pytorch-msssim. Updated November 17, 2021. 2023. https://github.com/jorge-pessoa/pytorch-msssim

40. Virtanen P, Gommers R, Oliphant TE, et al. SciPy 1.0: fundamental algorithms for scientific computing in Python. Nat Methods. Mar 2020;17(3):261–272. doi:10.1038/s41592-019-0686-2

41. Arapakis I, Efstathopoulos E, Tsitsia V, et al. Using “iDose4” iterative reconstruction algorithm in adults’ chest-abdomen-pelvis CT examinations: effect on image quality in relation to patient radiation exposure. Br J Radiol. Apr 2014;87(1036):20130613. doi:10.1259/bjr.20130613

